# Tau-PET in early cortical Alzheimer brain regions in relation to mild behavioral impairment in older adults with either normal cognition or mild cognitive impairment

**DOI:** 10.1101/2024.02.11.24302665

**Authors:** James Naude, Meng Wang, Rebeca Leon, Eric Smith, Zahinoor Ismail

**Author notes:** Corresponding Author: Zahinoor Ismail 3280 Hospital Dr. NW Calgary, Alberta, Canada T2N 4Z6.

## Abstract

Mild Behavioral Impairment (MBI) leverages later-life emergent and persistent neuropsychiatric symptoms (NPS) to identify a high-risk group for incident dementia. Phosphorylated tau (p-tau) is a hallmark biological manifestation of Alzheimer disease (AD). We investigated associations between MBI and tau accumulation in early-stage AD cortical regions. In 442 Alzheimer’s Disease Neuroimaging Initiative participants with normal cognition or mild cognitive impairment, MBI status was determined alongside corresponding p-tau and A*β*. Two meta-regions of interest were generated to represent Braak I and III neuropathological stages. Multivariable linear regression modelled the association between MBI as independent variable and tau tracer uptake as dependent variable. Among A*β* positive individuals, MBI was associated with tau uptake in Braak I (*β*=0.45(0.15), *p<.*01) and Braak III (*β*=0.24(0.07), *p<.*01) regions. In A*β* negative individuals, MBI was not associated with tau in the Braak I region (p=.11) with a negative association in Braak III (*p=.*01). These findings suggest MBI may be a sequela of neurodegeneration, and can be implemented as a cost-effective framework to help improve screening efficiency for AD.

## 1.0 Introduction

Alzheimer Disease (AD) is prevalent in 5.8 million Americans over the age of 65, characterized by progressive cognitive decline and functional impairment (Association, 2020). Current medications are mostly symptomatic treatments, and the search for disease modifying therapies has been challenging. From 2003 until recently, no clinical trial had met all primary endpoints, possibly due to intervention too late in the disease course (Gauthier et al., 2016; Mortby, M.E et al., 2018). Notwithstanding the recent success of the disease modifying therapy lecanemab and subsequent approval in the United States (Hoy, 2023), many other DMTs have failed, sometimes due to challenges recruiting participants with early-stage disease. The hallmarks of AD are Amyloid-*β* (A*β*) plaques and phospho-tau (p-tau) tangles; the bulk of research has focused on A*β*. However, tau is clinically meaningful, correlating better with cognitive symptoms, severity of dementia, and function, thus becoming an emerging focus of therapeutics (Congdon et al., 2023; Jack Jr et al., 2019; Malpas et al., 2020). Detecting prodromal and preclinical disease often requires neuropsychological testing, followed by ligand-based PET imaging or CSF biomarker analyses for AD biomarker confirmation, although plasma biomarkers are gaining traction (Balogun et al., 2023). Nonetheless, barriers remain to widespread screening for plasma biomarkers, including access for those in remote geographies, high cost of assays on top of blood draws, and the risk of false positives. In clinical trials, biomarkers are costly and inefficient if used to screen for AD+ cases. Neuropsychiatric symptoms (NPS) may offer an inexpensive and efficient opportunity for trial enrichment for biomarker positivity. If applied before biomarker assays, adding NPS assessments may improve accuracy of risk estimates generated from cognitive symptoms alone (especially in preclinical AD), identifying a sub-group with substantially higher risk, therefore increasing screening yield and detection of disease (Soto et al., 2023).

Historically overshadowed by cognitive symptoms in AD, NPS have emerged as a key component of the disease. NPS are almost ubiquitous in dementia (Lanctôt et al., 2017), but are also common in mild cognitive impairment (MCI) where symptoms are associated with faster progression to dementia (Martin and Velayudhan, 2020). NPS emerge in advance of cognitive symptoms in 59% of all-cause dementia, including 30% of those who develop AD (Wise et al., 2019). In cognitively normal (NC) older adults, the presence of NPS has been associated with cognitive decline and dementia (Burhanullah et al., 2019; Geda et al., 2014; Liew, 2020). However, the assessment of NPS in older adults can be challenging; in many cases psychiatric diagnoses are provided, and neurodegenerative disease is not initially considered on the differential diagnosis (Cieslak et al., 2018; Matsuoka et al., 2019; Mortby et al., 2017).

Mild Behavioral Impairment (MBI) is a validated dementia risk syndrome (Ismail et al., 2016) distinct from chronic and/or recurrent psychiatric illness (Matsuoka et al., 2019; Taragano et al., 2018). While conventionally measured NPS are associated with incident dementia, MBI leverages neurodegenerative disease associations with later-life emergent and persistent NPS to identify a group at much higher risk (Bateman et al., 2020; Creese et al., 2023; Creese et al., 2019; Ebrahim et al., 2023; Gill et al., 2020; Gill, Sascha et al., 2021; Ismail et al., 2023a; Ismail et al., 2021; Kan et al., 2022; Matsuoka et al., 2019; McGirr, A. et al., 2022; Rouse et al., 2023; Vellone et al., 2022; Yoon et al., 2022). Several recent papers have demonstrated this point when MBI was compared to psychiatric disorders or NPS not meeting MBI criteria, with the MBI group having faster cognitive decline and progressing more rapidly to dementia (Ebrahim et al., 2023; Ghahremani, Maryam et al., 2023; Ismail et al., 2023a; Ismail et al., 2023b; Matsuoka et al., 2019; Taragano et al., 2018; Vellone et al., 2022), and even lower reversion rates from MCI to NC (McGirr, Alexander et al., 2022). For some, MBI is a proxy marker of underlying neurodegenerative disease pathology, associated with AD risk genes, amyloid, tau, and neurodegeneration (Andrews et al., 2018; Creese et al., 2021; Ghahremani, Maryam et al., 2023; Gill, S. et al., 2021; Ismail et al., 2023b; Johansson et al., 2021; Lussier et al., 2020; Matsuoka et al., 2023; Matsuoka et al., 2021; Matuskova et al., 2021; Miao et al., 2021a; Naude et al., 2020). Thus, a greater proportion of the MBI group has underlying AD compared to the conventional psychiatric/NPS group; this represents prodromal AD for MBI in MCI, and preclinical AD for MBI in NC. These data tell us that risk estimates based on the emergence and persistence of NPS are useful, and that MBI should be reported in conjunction with cognitive status, contributing an estimate of behavioral risk as a complement to the determination of risk based on cognition.

Much of the MBI biomarker literature has focused on fluid biomarkers and structural (Matsuoka et al., 2023) and functional (Ghahremani, M. et al., 2023) imaging. Relatively little data have explored ligand-based PET imaging. PET studies are important to determine locations and patterns of amyloid and tau binding, to explore how presentation of MBI aligns with known patterns in AD. The Canadian TRIAD study first explored this concept in 96 NC participants, finding a correlation between MBI Checklist (MBI-C) (Ismail et al., 2017) score and global and striatal A*β*-PET tracer uptake was shown; however, the same was not demonstrated for tau-PET tracer (Lussier et al., 2020). Subsequently, a Swedish Biofinder2 study explored this same relationship in a sample of 50 A*β+* NC participants, finding an association between MBI-C score and tau-PET in the entorhinal cortex and hippocampus (Johansson et al., 2021). These conflicting results necessitate further exploration of the association of tau-PET with MBI in the presence of A*β*. Here, in a sample of NC/MCI individuals including both A*β+* and A*β-* participants, we aimed to determine whether persons with NPS had tau-PET tracer uptake in regions of the brain affected early in the course of AD. We hypothesized that participants with a NPS profile consistent with MBI, would have a greater standardized uptake value ratio (SUVR) of AV1451 (flortaucipir), compared with persons with NPS profiles not consistent with MBI.

## 2.0 Methods and Materials

### 2.1 Alzheimer’s Disease Neuroimaging Initiative (ADNI)

Data are from the Alzheimer’s Disease Neuroimaging Initiative (ADNI), a partnership involving multiple centers across North America with the goal of tracking participants through periods of cognitive decline and dementia. Launched in 2003, ADNI continues to evaluate biomarker, neuroimaging, and neuropsychological status in participants. Informed consent was obtained from all participants prior to enrolling in the study. Data in this study were collected prior to August 2022.

### 2.2 Sample

Participants enrolled in ADNI with AV1451 tau-PET and AV45 amyloid-PET imaging data available were included (n=442). Participants were classified as either NC or MCI at the time of tau-PET imaging. Participants free of memory complaints, scoring between 24-30 on the MMSE, with a Clinical Dementia Rating (CDR) of 0, and a Memory Box Score of 0 were NC. MCI was diagnosed if there were subjective memory concerns, MMSE between 24-30, CDR of 0.5, and Memory Box Score ≥0.5. Participants were excluded from this study if at the imaging visit NPS score was not available for either the visit associated with tau-PET or the subsequent visit, A*β*-PET status was not determined, or diagnosis was dementia. Demographic information including age, sex, and years of education were obtained at the time of the AV1451 imaging visit. Detailed participant selection is displayed in Figure 1.

**Figure 1:**
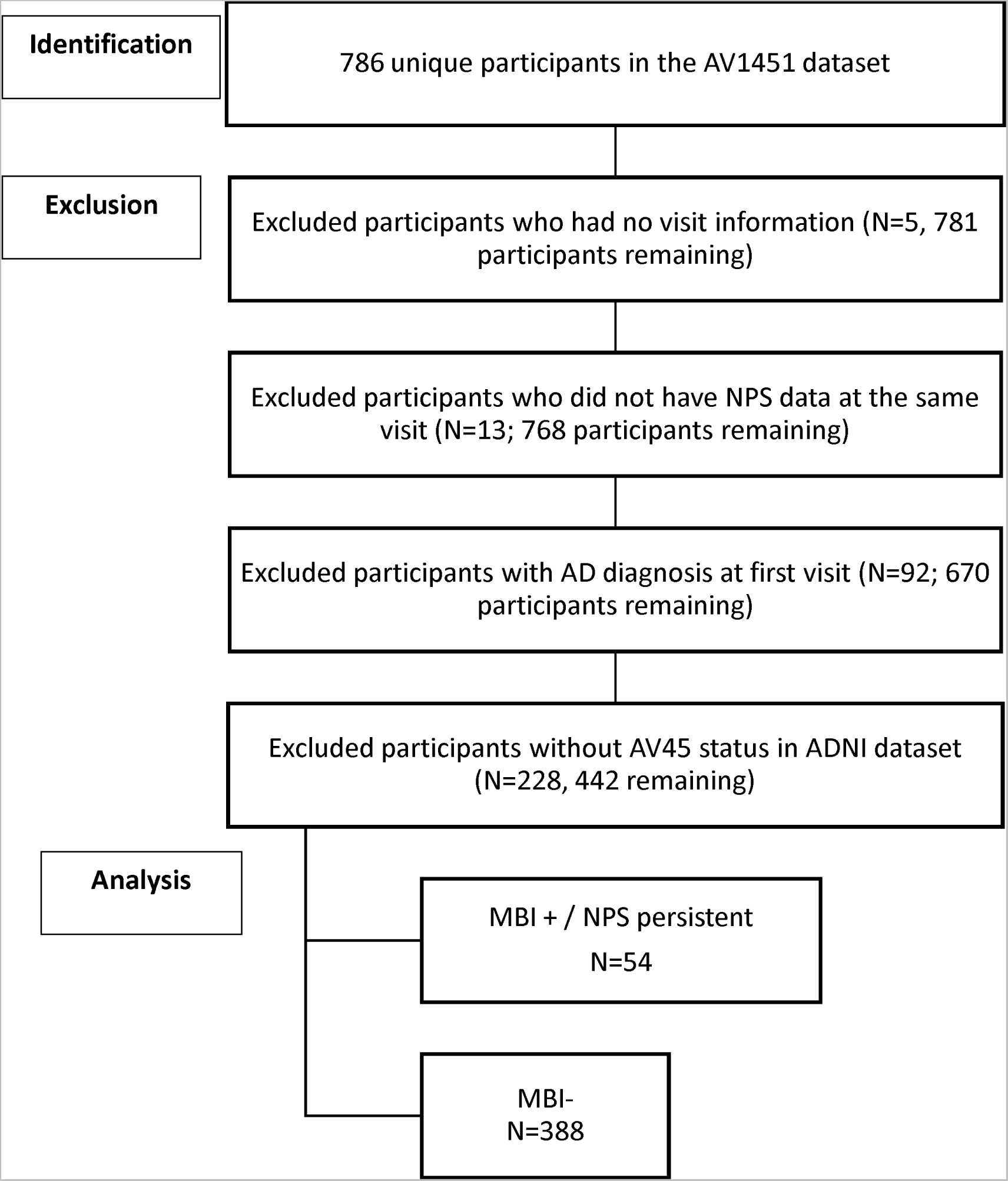
Depicts derivation of the sample population from ADNI datasets.

### 2.3 NPS Case Ascertainment and MBI Status

The Neuropsychiatric Inventory (NPI) (Cummings, 2020) was used to generate a NPS score that mapped NPI items onto MBI domains (Mortby, Moyra E et al., 2018; Sheikh et al., 2018). The NPI, clinician-rated after informant interview, consists of 12 items relating to frequency and severity of NPS over the previous month. The 5 MBI domains were derived from 10 NPI domains as follows: MBI motivation/drive from NPI apathy/indifference; MBI emotional regulation from the sum of NPI depression, anxiety, and elation/euphoria; MBI impulse control from the sum of NPI irritability, agitation/aggression, and aberrant motor behavior; MBI social cognition from NPI disinhibition; and MBI thoughts/perception from NPI delusions/hallucinations. Neurovegetative NPI items for sleep and appetite were not included as neither map well onto MBI criteria. If score=0 on all selected NPI items, the visit was coded NPS-. If score was >0 on any of the select NPI domains, the visit was coded NPS+. MBI status was determined from NPS status at the tau-PET scan visit and the visit 6 months later (Guan et al., 2023). A two-level variable for MBI was utilized (MBI+, MBI-), informed by recent ADNI data demonstrating that MBI was associated with significantly higher p-tau levels relative to no NPS, but NPS not meeting MBI criteria did not differ from the no NPS group (Ghahremani, Maryam et al., 2023). Thus, NPS+ status at both visits (symptom persistence) was classified as MBI+, and NPS-status at either or both visits was classified as MBI-. If NPI data were available for only one visit, the participant was conservatively classified as MBI-.

### 2.4 AV1451 and Regions Included

Partial Volume Corrected (PVC) AV1451 PET data were processed to render standardized uptake values (SUVs), which were reported for left and right hemispheric components of structures. Each regional SUV was normalized by dividing the raw SUV by the inferior cerebellar gray matter SUV to generate a ratio, the SUVR. Left and right hemispheric components of structures were summed and meaned. Brain regions affected early in the AD course were chosen based on previous research on tau binding and spread pattern (Cho et al., 2016; Jack et al., 2018; Ossenkoppele et al., 2018). Two meta-regions of interest (ROIs) representing early Braak neuropathological stages (Braak and Braak, 1991) were generated for use in the analysis. The Braak neuropathological staging system is based on the distribution of neurofibrillary tangles (NFTs), composed of tau. According to the staging system, NFTs are first seen in the entorhinal cortex, then hippocampus, and then other regions (Braak and Braak, 1991). For our analysis, the first ROI represented Braak stage I, composed of both left and right entorhinal cortex. The second ROI was generated to represent Braak III, composed of bihemispheric components of the parahippocampus, fusiform, posterior cingulate gyrus, lingual gyrus, and amygdala. A ROI was not generated for the hippocampus, representing Braak stage II, due to substantial off-target binding of AV1451 (Lee et al., 2018). A*β+* status was determined based on AV45 SUVR ≤1.11 (Landau et al., 2012). Further information on the image processing pipeline for this study is available at adni.loni.usc.edu.

### 2.5 Statistical Analysis

Sample characteristics were described using mean, standard deviation, quartile, and frequency distributions. To assess the relationship between MBI status and AV1451 tau-PET SUVR for both ROIs, multivariable linear regression models were fitted with MBI status as the independent variable and tau-PET SUVR as dependent variable, adjusting for age, sex, and education years, with testing of interaction terms for MBI*cognitive status (NC/MCI) and MBI*A*β* status (+/-). The assumptions of linear regression were confirmed by examination of the residuals. All results were considered statistically significant with a two-sided *p*-value of <0.05. Analyses were conducted in SAS (v9.4 SAS Institute Inc., 2013).

### 2.6 Data availability

Data used in this study are available from adni.loni.usc.edu for download.

## 3.0 Results

Participant mean age was 75 years (SD=7.6), 64% had normal cognition, and about half were females (50.9%), with median educational attainment of 16 years (Table 1). Of the 442 participants, 338 were MBI- and 54 MBI+, 252 A*β*- and 188 A*β*+, 283 NC and 157 MCI (Table 2).

**Table 1.**
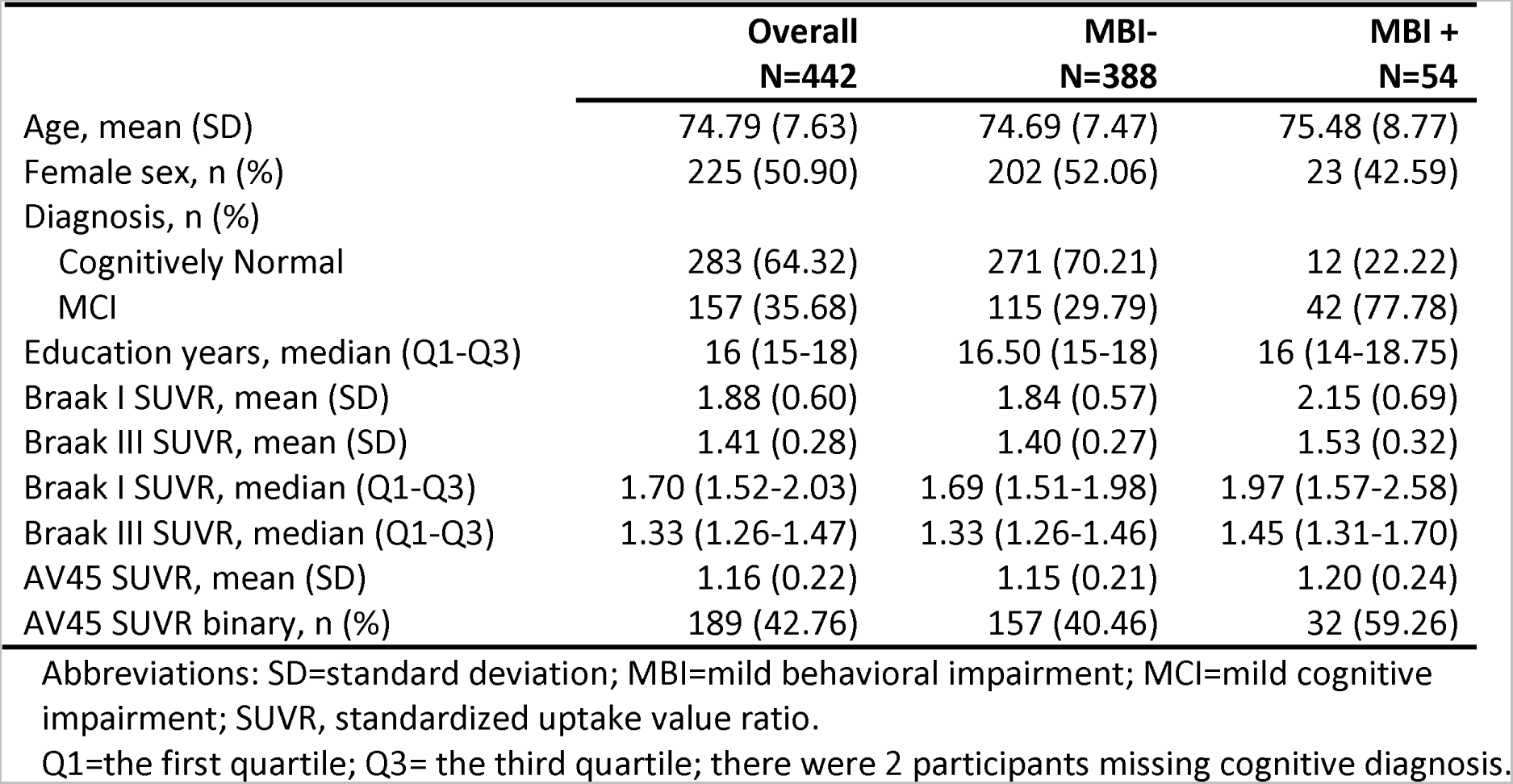
Sample Characteristics for overall sample and by MBI status.

**Table 2.**
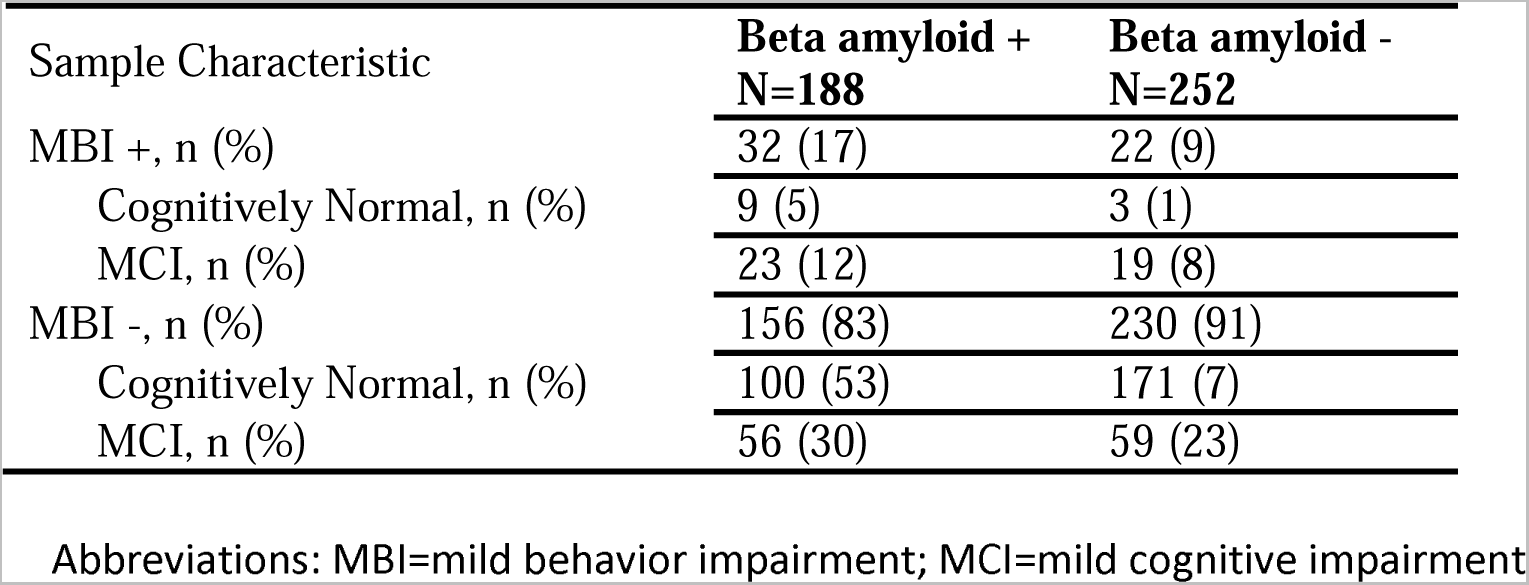
Cognitive and MBI characteristics stratified by amyloid status.

In the ROIs representing Braak I and Braak III neuropathological stages, A*β* status moderated the relationship between MBI status and AV1451 SUVR, after controlling for age, sex, education, and cognitive diagnosis (Table 3). Specifically, MBI*A*β* interaction coefficients were 0.45 (95%CI: 0.16-0.75, p< 0.01) for the Braak I ROI, and 0.24 (95%CI: 0.11-0.38, p<0.01) for the Braak III ROI. In the A*β+* stratum, MBI was significantly associated with greater AV1451 SUVR in the Braak I ROI (*β̂*=0.26, 95%CI: 0.07-0.46, p<0.01) and the Braak III ROI (*β̂*=0.11, 95%CI: 0.02-0.20, p=0.01). In the A*β-* stratum, MBI status was not associated with AV1451 SUVR in the Braak I ROI (*β̂* =-0.19, 95%CI: -0.42 to 0.04, p=0.11) and was negatively associated with SUVR in the Braak III ROI (*β̂*=-0.13, 95%CI: -0.24 to -0.03, p=0.01). The adjusted marginal means of tau SUVR with 95% CI stratified by MBI status and A*β* status are shown in Figure 2.

**Figure 2.**
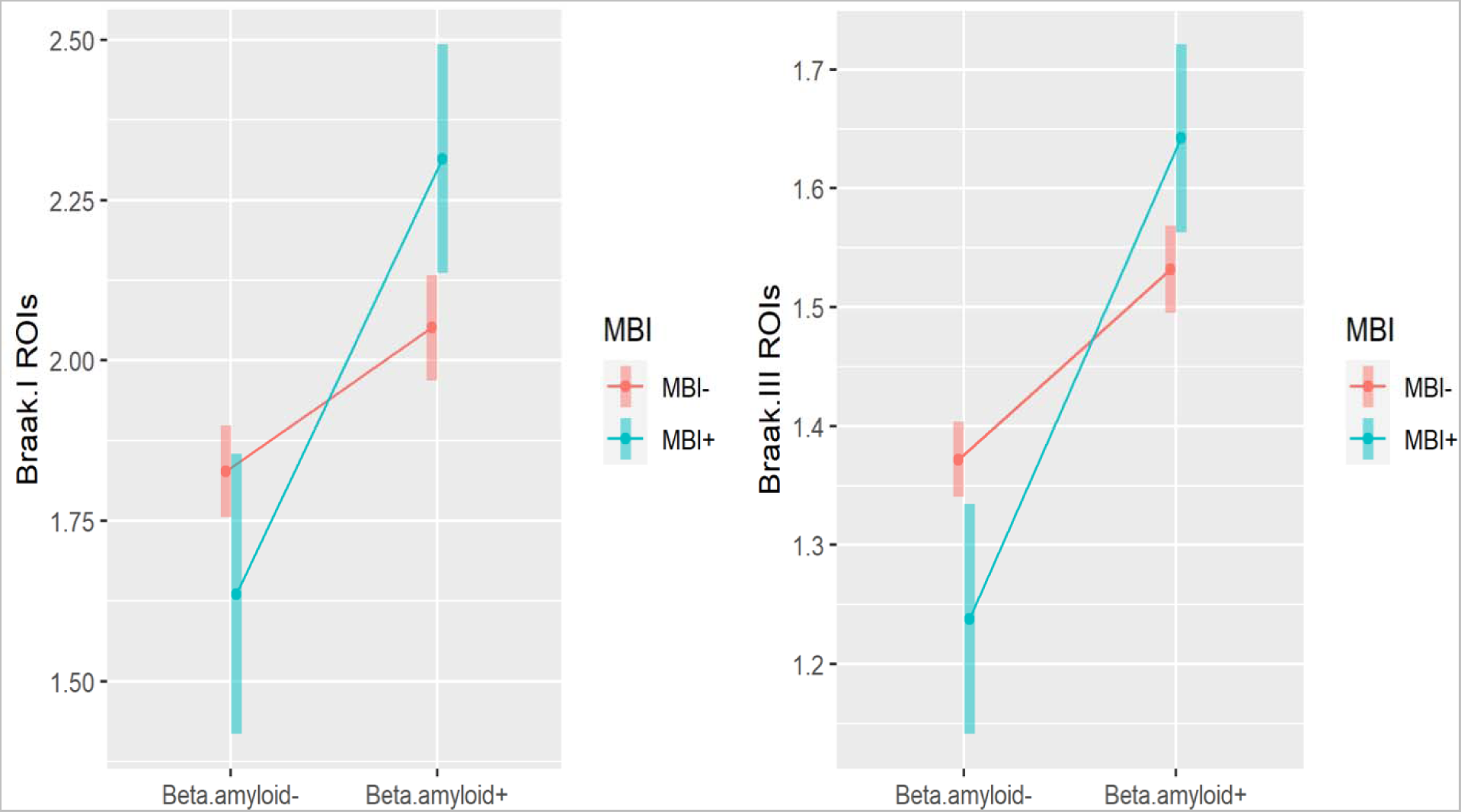
Association of MBI with TAU (Braak I and Braak III), by beta-amyloid status. Plots of the marginal means of tau with 95% CI stratified by MBI status and beta-amyloid status are based on multiple linear regression, for Braak I and Braak III, respectively.

**Table 3.** The association between MBI and Braak I and III SUVR adjusted for age, sex, education, MCI, and AV45 using multiple linear regression models.

|  | Outcome variable<br>Braak I tau SUVR<br>N=440 |  | Outcome variable<br>Braak III tau SUVR<br>N=440 |  |
| --- | --- | --- | --- | --- |
| | $\hat{\beta}$ (95%CI) | *p* value | $\hat{\beta}$ (95%CI) | *p* value |
| **Independent Variables:** |  |  |  |  |
| **Aβ +** |  |  |  |  |
| **MBI + vs MBI -** | 0.26 (0.07, 0.46) | <0.01 | 0.11 (0.02, 0.20) | 0.01 |
| **Aβ -** |  |  |  |  |
| **MBI + vs MBI -** | -0.19 (-0.42, 0.04) | 0.11 | -0.13 (-0.24, -0.03) | 0.01 |
| MCI vs normal cognition | 0.44 (0.34, 0.55) | <0.01 | 0.22 (0.17, 0.26) | <0.01 |
| Age | 0.002 (-0.004, 0.01) | 0.45 | 0.002 (-0.0004, 0.01) | 0.10 |
| Female vs male | -0.10 (-0.20, 0.003) | 0.06 | -0.02 (-0.07, 0.02) | 0.32 |
| Education years | 0.01 (-0.01, 0.03) | 0.28 | 0.005 (-0.003, 0.01) | 0.23 |

Abbreviations: SD=standard deviation; MBI=mild behavioral impairment; MCI=mild cognitive impairment; SUVR, standardized uptake value ratio. SE= standard error. $\hat{\beta}$ is the estimated regression coefficient.

MCI cognitive status was associated with higher AV1451 SUVR, in both the Braak I ROI (*β̂*=0.44, 95%CI: 0.34-0.55, p<0.01) and the Braak III ROI (*β̂*=0.22, 95%CI:0.17-0.26, p<0.01), but significant interaction effects between cognitive status and MBI were not found (p=0.24). Age, sex, and education were not associated with AV1451 SUVR in either ROI.

## 4.0 Discussion

In this relatively large study of 442 dementia-free participants, the majority of whom were cognitively normal, A*β* status moderated the association between MBI and tau-PET SUVR in the ROIs representing Braak I and Braak III stages of AD progression. In the A*β*+ stratum, when compared to the group with no NPS or NPS not meeting the MBI symptom persistence criterion, participants with MBI had higher tau tracer uptake in the Braak I and Braak III ROIs adjusted for age, sex, education, and cognitive diagnosis. In the A*β*-stratum, MBI was not significantly associated with tau uptake in the Braak I ROI and had a negative association with SUVR in Braak III. These findings suggest that AD and not minor age-related tauopathy (Bell et al., 2019) may be the main driver of MBI.

### 4.1 Braak Stages

Our representation of Braak I consisted of a weighted mean of the left and right entorhinal cortices only, normalized to the inferior cerebellar grey matter consistent with other ADNI studies (Yasuno et al., 2020). Tauopathies first become apparent within the entorhinal cortex during AD pathological progression (Cho et al., 2016), advancing to the hippocampus and then other temporal lobe structures. Entorhinal tau accumulation was consistent with our hypothesis. A post-mortem study presented compatible findings, with NPI-derived NPS such as anxiety, agitation, and depression associating with tau pathology and neurofibrillary tangles in Braak I and II regions (Ehrenberg et al., 2018). These findings suggest that both cognitive and behavioral manifestations of AD relate to p-tau neurofibrillary tangles in the entorhinal cortex, supporting the notion that NPS are core AD features. Due to technical constraints associated with off-target radiotracer binding, however, the hippocampal regions were not included in this study. The Johansson (2021) study, however, did find MBI associations with hippocampal tau SUVR. As Braak III is a stage of tau accumulation downstream from Braak stages I and II, our findings support MBI as a behavioral manifestation of AD that follows a classical progression of tau pathology (Braak et al., 2006; Davidson et al., 2018; Vogel et al., 2021).

### 4.2 Tau

Accumulation of A*β* pathology is considered the initial event in the AD pathological process, long before clinical symptoms (Jack Jr et al., 2019) with several MBI studies demonstrating MBI associations. Tau accrues temporally downstream from A*β* (Gordon and Tijms, 2019); abnormal tau-PET rarely occurs in the absence of abnormal A*β*-PET, with the tauopathy associating with clinical symptoms (Jack Jr et al., 2019). The tau protein stabilizes the microtubule network to ensure proper neuronal function and communication (Iqbal et al., 2010). In AD, tau becomes hyperphosphorylated, leading to production of neurofibrillary tangles, preventing normal cytoskeletal activities of microtubule associated proteins, which results in cognitive symptoms (Iqbal et al., 2010; Jack Jr et al., 2019). Post-mortem studies have determined that tau is the best predictor of global cognitive status, clinical dementia stage, functional abilities, and NPS (Malpas et al., 2020). In persons with dementia, or mixed samples including dementia, some studies have demonstrated associations between tau and NPS (Banning et al., 2020; Bloniecki et al., 2014; Koppel et al., 2013; Tommasi et al., 2021; Yasuno et al., 2020). In persons without dementia, some small to mid-size studies have found associations between tau and NPS (Ng et al., 2021). Tau has been linked with affective symptoms (Babulal et al., 2020; Gatchel et al., 2017; Yasuno et al., 2020), anxiety (Ramakers et al., 2013), and apathy (Binette et al., 2020; Johansson et al., 2022). However, there have also been a number of studies that failed to find associations between tau and overall NPS burden (Binette et al., 2020) or specific NPS domains including agitation, anxiety, apathy, depression, hyperactivity, irritability, and psychosis (Auning et al., 2015; Binette et al., 2020; Johansson et al., 2022; Ramakers et al., 2013; Yasuno et al., 2020). Interestingly, in one study of NC participants, no baseline associations between CSF p-tau181 and NPS were found, but p-tau was associated with an increase in NPS at 1 year (Babulal et al., 2016). These findings suggest that tau may be associated with incident or worsening NPS, consistent with the findings of our study.

Few studies have explored the relationship between tau and MBI specifically. BioFINDER2 assessed tau-PET [18F]RO948 retention and CSF P-tau181 levels in 50 NC A*β* positive participants, i.e., with preclinical AD or Stage 2 disease as per the NIA-AA framework (Knopman et al., 2018). The MBI-C was used to measure MBI symptoms. Regression models determined that MBI-C score was associated with higher tau-PET SUVR in a Braak I-II composite ROI (but not Braak III-IV or V-VI). MBI was also associated with significantly higher CSF p-tau. In this NC sample, MBI-C score but not memory deficits predicted PET and CSF-tau, emphasizing the importance of MBI in advance of objective cognitive changes (Johansson et al., 2021). Our findings are consistent, demonstrating an association between MBI and tau, in A*β*+ participants with preclinical (NC) and prodromal (MCI) AD. TRIAD assessed the relationship between tau-PET [18F]MK6240 and MBI, also measured with the MBI-C, in 96 NC participants and 91 with MCI. In the NC group, while there were significant positive correlations between MBI-C score and global and striatal A*β*-PET [18F]AZD4694 SUVR, no significant associations were found between MBI and Braak Stage I or II tau-PET binding (Lussier et al., 2020). From a preliminary analysis in the MCI group, however, MBI was associated with tau signal in early AD regions particularly the precuneus and posterior cingulate cortex bilaterally (Lussier et al., 2019). We speculate that there were insufficient A*β*+ positive participants in the TRIAD NC sample to drive a tau signal, but sufficient numbers in the MCI sample to find a MBI relationship with tau.

In contrast, a recent ADNI study showed an association between MBI symptoms and A*β*-PET AV45, but no association between MBI and tau-PET AV1451 (Sun et al., 2021). Of note, a single-visit approach was taken in this study, mapping NPS onto MBI criteria, but without consideration of symptom persistence. Symptom persistence is a core MBI criterion, associated with greater progression from MCI to dementia, and lower reversion from MCI to NC (McGirr, A. et al., 2022). In our study, a complementary approach was taken, requiring NPS persistence over two study visits for MBI+ status. Consistent with the signals in BioFINDER2 and TRIAD, individuals with MBI had significantly greater burden of tau pathology in regions affected early in the course of AD, compared to the group without MBI. This finding supports the need to incorporate natural history into assessment of NPS in older adults, even those without cognitive symptoms. Rather than a cross-sectional measure of NPS at one single time point, the foundation of the MBI framework lies in assessing the natural history of NPS. Symptom *emergence in later in life* is a fundamental feature to help differentiate MBI from chronic and recurring psychiatric conditions. Symptom *persistence* differentiates MBI from transient and reactive NPS, which are less likely to represent behavioral sequelae of neurodegenerative disease than MBI. Thus MBI improves specificity and signal-to-noise ratio for disease detection. A very recent study of MBI and plasma p-tau181 has further extended this clinical approach. In a sample of 571 dementia-free participants in ADNI, MBI was significantly associated with higher p-tau181 levels compared to groups with NPS not meeting MBI criteria, and with no NPS. Longitudinally, MBI was associated with increasing p-tau, declining memory and executive function, and a 3.92-fold greater incidence rate of dementia (Ghahremani, Maryam et al., 2023). Similarly, in a study of MCI participants from ADNI and MEMENTO, MBI was associated with higher CSF p-tau levels at baseline, and increasing CSF p-tau levels over 4 years compared to the group with NPS not meeting MBI criteria (Ismail et al., 2023b). This finding was consistent in the ADNI test cohort (observational cohort) and in the MEMENTO validation cohort (memory clinic patients). In sum, this small group of novel studies are consistent in finding that MBI is associated with greater A*β*-related tau burden, supporting the biological underpinnings of MBI, and its role in capturing early-stage AD.

In our study, it was also important to resolve the role of A*β* status on the relationship between MBI and tau, given the conflicting findings in the Lussier and Johansson studies (Johansson et al., 2021; Lussier et al., 2020). MBI describes an elevated AD risk state and A*β* indicates the beginning of significant disease burden, usually preceding tau pathology (Gordon and Tijms, 2019). Thus, our findings that those with both A*β* and MBI positivity showed the greatest tau burden was unsurprising. Exploration of the A*β* negative stratum, however, was particularly informative. In both the Braak I and III ROIs, MBI positivity was negatively associated with tau-PET SUVR, although this effect was only statistically significant in Braak III regions. While these results might appear counterintuitive at first glance, they support the link between MBI and core early AD pathology, i.e., A*β*, without which there is little tau (Jack Jr et al., 2019). AV1451 binds paired helical filament tau, which is a combination of 3R and 4R tau. In contrast, for A*β* negative participants with MBI, other pathology may be involved, including vascular pathology, alpha synuclein, TDP-43, and non-AD tauopathies, e.g., straight or twisted non-helical tau, or just 3R or 4R tau, rather than the combination (Gibson et al., 2022; Mattsson et al., 2019; Miao et al., 2021b; Tsai et al., 2019). Future studies in A*β* negative participants exploring associations between MBI and other pathologies will help clarify this issue as well as explore MBI specifically in non-AD populations, once technology progresses enough to reasonably allow incorporation of fluid biomarkers and imaging tracers that bind these other pathological proteins.

Nonetheless, our study findings suggest that this MBI approach to NPS case assessment can help identify older adults, with at most MCI, at the earliest phase of cortical tauopathy due to AD. The results also lend credence to longitudinal studies finding that MBI is associated with faster cognitive decline (Creese et al., 2019), and a greater risk of developing AD (Gill et al., 2020). In a recent clinicopathological study of NACC participants, MBI in cognitively normal older adults was associated with a higher hazard of both clinically-diagnosed, and neuropathologically-confirmed AD over the next 5 years (Ruthirakuhan et al., 2022). More recently, in a functional MRI study, MBI was associated with connectivity changes in the Default Mode and Salience Networks typically seen in early-stage AD (Ghahremani, M. et al., 2023). These findings suggest a neurobiological basis of behavioral and psychological symptoms in this dementia-free sample of older adults, specifically suggesting an association to MAP systems and cytoskeletal components of neurons in varying regions of the brain (Dehmelt and Halpain, 2004). Another study suggested that cytoskeletal components such as neurofilament light were important in the presenting symptoms of MBI, and especially the emergence of those MBI symptoms, adding further credence to this approach (Naude et al., 2020).

Assessment of MBI is an effective approach that can be used in conjunction with cognitive status to identify individuals at greater risk of cognitive decline. With tau therapeutic agents on the horizon (Congdon et al., 2023) these findings may be of particular interest. The emergence and persistence of MBI symptoms might serve as a proxy marker of entorhinal tau pathology, which could be considered in clinical screening and for clinical trials (Mortby, M.E et al., 2018). For example, in dementia-free older adults, assessment of MBI could be scaled up and implemented remotely (Creese et al., 2020; Kassam et al., 2023), to flag a group for further clinical and biomarker workup. This initial remote pre-screen with an informant-rated measure of MBI could increase efficiency for preclinical or prodromal case detection and reduce costs in clinical trials due to a reduction in screen failures. Further, MBI may serve as a treatment target (Soto et al., 2023), to either reduce symptom burden, or as part of a multidimensional collection of symptoms identified in the meaningful benefits framework, to evaluate effects of disease-modifying therapies (Assunção et al., 2022).

### 4.3 Limitations

Despite the novelty of the study and the substantial sample size, several limitations are important to consider. The AV1451 radiotracer limited assessment of Braak Stage II due to offsite choroid plexus binding (Lee et al., 2018). Future studies examining MBI and tauopathy will need to establish whether there exist any relationships within this region. Braak III was represented as a meta-ROI consisting of several regions, which might result signal loss in some regions within the composite. However, due to multicollinearity, we did not assess individual regions separately. The gold standard for MBI case ascertainment is the MBI-C, which is not utilized in ADNI. Although we approximated MBI status, transforming NPI items using a validated algorithm, this approach may not be as sensitive or specific in capturing MBI (Hu et al., 2023; Mallo et al., 2018; Mallo et al., 2019). For example, the persistent NPS group may have had emergence earlier in ADNI, before tau PET was implemented, or may have longstanding subsyndromal psychiatric symptoms, not screened out at study entry. Utilizing the MBI-C in future studies may resolve these issues and provide more clarity. Additionally, in A*β* PET studies of neurodegenerative disease, lack of stratification by apolipoprotein IZ4 (APOE4) status has been cited as a potential contributor to inconclusive findings (Oliveira, 2023). This logic likely applies to tau-PET studies as well. Unfortunately, APOE4 data were not available for all participants; using this reduced sample would have resulted in a significant loss of statistical power for the interaction analyses. Nonetheless, addressing APOE4 effects is an important issue for studies of cognition, behavior, and function, and future studies should incorporate APOE as a covariate in the analysis. Furthermore, we were unable to assess longitudinal associations between MBI and tau tangle burden, as the implementation of tau-PET in ADNI is a relatively recent advance with insufficient sample size for analysis. By assessing these trajectories in the future, we can elucidate the differing rates of tau accumulation between those who meet MBI criteria and those who do not.

### 4.4 Conclusions

In this sample of dementia-free participants, the majority of whom were cognitively normal, MBI was associated with greater tau neuropathological burden in early cortical regions affected in AD, in A*β* positive participants. The findings support the biological understanding of NPS as part of the AD process, detectable at preclinical and prodromal stages. This study adds further support to the relevance of the MBI construct for dementia detection and prognostication. Future studies can explore MBI as an approach to clinical trial enrichment, to improve screening efficiency for A*β* and/or tau positivity, and to decrease study cost and duration for interventions targeting these proteins.

## Data Availability

All data produced in the present study are available upon reasonable request to the Alzheimer's Disease Neuroimaging Initiative.

## 5.0 Acknowledgements

Data used in preparation of this article were obtained from the Alzheimer’s Disease Neuroimaging Initiative (ADNI) database (http://adni.loni.usc.edu). As such, the investigators within the ADNI contributed to the design and implementation of ADNI and/or provided data but did not participate in analysis or writing of this report. A complete listing of ADNI investigators can be found at: http://adni.loni.usc.edu/wpcontent/uploads/how_to_apply/ADNI_Acknowledgement_List.pdf. Data collection and sharing for this project was funded by the Alzheimer’s Disease Neuroimaging Initiative (ADNI) (National Institutes of Health Grant U01 AG024904) and DOD ADNI (Department of Defense award number W81XWH-12-2-0012). ADNI is funded by the National Institute on Aging, the National Institute of Biomedical Imaging and Bioengineering, and through generous contributions of several others.

Z.I. is funded by the Canadian Institutes of Health BCA2633. This study was also supported by the UK National Institute for Health and Care Research Exeter Biomedical Research Centre. The views expressed are those of the author(s) and not necessarily those of the NIHR or the Department of Health and Social Care.

